# Efficacy and Safety of Esketamine Nasal Spray in Treatment-Resistant Depression: A Systematic Review and Meta-Analysis of Randomized Controlled Trials

**DOI:** 10.1101/2025.09.22.25336317

**Authors:** Bhaskara Gowthami, Ishani Mehta, Sneha Baiju, Ashish Appani, Tanisha Suvarna, Kiratjyot Singh Randhawa, Anuj Manish Kakkad, Afzal Biabani Syed, Vaishnavi Kumar, Harshawardhan Dhanraj Ramteke, Rakhshanda khan

## Abstract

**Introduction:** Treatment-resistant depression (TRD) affects up to one-third of patients with major depressive disorder, leading to poor outcomes and increased suicide risk. Esketamine nasal spray, a novel glutamatergic modulator, has emerged as an adjunctive option with rapid onset of action. However, the efficacy and safety of esketamine across randomized controlled trials (RCTs) remain variably reported, necessitating a systematic synthesis.

**Methods:** We systematically searched PubMed, Embase, Cochrane CENTRAL, Web of Science, and ClinicalTrials.gov from inception to September 2025. Seventeen RCTs comprising 10,073 patients were included, of whom 5,707 received esketamine plus oral antidepressant and 4,622 received placebo plus oral antidepressant. Primary efficacy outcomes included change in depressive symptoms, response (≥50% reduction), and remission rates. Secondary outcomes were functional improvement (Sheehan Disability Scale, SDS) and safety events (dissociation, sedation, hypertension, nausea). Random-effects models were used to pool mean differences (MD), odds ratios (OR), and risk ratios (RR) with 95% confidence intervals (CI).

**Results:** Esketamine significantly improved response (OR = 0.51; 95% CI: 0.30–0.73; p < 0.001; 14 RCTs) and remission (OR = 0.35; 95% CI: 0.11–0.58; p < 0.01; 13 RCTs). Functional outcomes also favored esketamine (SDS MD = –2.27; 95% CI: –3.50 to –1.04; p < 0.01; 4 RCTs). Pooled analysis of continuous MADRS and CGI-S change showed non-significant differences (MADRS MD = –1.47; 95% CI: –3.01 to 0.07; CGI-S MD = –0.30; 95% CI: –0.75 to 0.14). Safety analysis revealed increased risk of dissociation (RR = 1.98; 95% CI: 1.68–2.28; 9 RCTs) and hypertension (RR = 1.42; 95% CI: 1.04–1.80; 9 RCTs), with non-significant elevations for sedation (RR = 1.23; 95% CI: 0.80–1.66) and nausea (RR = 1.10; 95% CI: 0.82–1.37).

**Conclusion:** Esketamine nasal spray plus oral antidepressant significantly improves treatment response, remission, and functioning in TRD patients but is associated with increased risk of dissociation and hypertension. While efficacy is robust, safety monitoring and structured clinical delivery remain essential. Further long-term and comparative effectiveness studies are warranted to define esketamine’s role in TRD management.

## Introduction

Major depressive disorder (MDD) is a leading cause of disability globally, imposing substantial psychological, social, and economic burdens [1]. Despite the availability of multiple established treatments, a significant proportion of patients fail to achieve adequate response, remission, or functional recovery. When depressive symptoms persist despite trials of two or more antidepressants at adequate dose and duration, the condition is commonly termed **treatment-resistant depression** (TRD). TRD is associated with increased morbidity, risk of suicide, poorer quality of life, and higher healthcare utilization. Rapid and robust treatment options are thus urgently needed for this group [2].

Ketamine, an N-methyl-D-aspartate (NMDA) receptor antagonist, has drawn considerable interest for its rapid antidepressant effects. However, the intravenous form is logistically burdensome, and its psychotomimetic and dissociative side effects, as well as potential for abuse, limit wider use [3]. Esketamine, the S-enantiomer of ketamine, has greater potency at NMDA receptors, promising similar antidepressant efficacy with potential advantages in dosing, tolerability, and delivery [4]. Intranasal esketamine has been developed and approved (in many jurisdictions) as an adjunctive therapy with oral antidepressants for TRD, with evidence of more rapid onset of action than conventional antidepressants.

Key randomized controlled trials (RCTs), including the TRANSFORM and SUSTAIN programs, demonstrated that esketamine nasal spray plus an oral antidepressant yields significantly greater reductions in depressive symptom severity (often measured via the Montgomery-Åsberg Depression Rating Scale, MADRS), higher response rates (typically ≥50% reduction in MADRS score), and improved remission rates (MADRS below a cut-off) compared to placebo plus antidepressant [5,6]. These trials also evaluated the trajectory of improvement, showing effects as early as 24 hours post-dose in some studies. Moreover, assessments of functional outcomes (e.g., using the Sheehan Disability Scale) and quality of life have provided complementary evidence that symptom improvement translates into real-world benefits. The safety/tolerability profile has been characterized by transient dissociation, sedation, increases in blood pressure, and common adverse effects such as nausea, dizziness, etc., generally manageable under supervised use. Long-term studies (e.g., up to 1 year in open-label extensions) have sought to clarify durability of efficacy and safety under more naturalistic conditions [7].

Despite a growing evidence base, several important gaps and controversies remain. First, the magnitude and consistency of treatment effects across different RCTs vary, especially for outcomes such as remission and functional recovery. Some trials report strong early onset but diminishing incremental benefits at longer follow-up; others show high placebo-adjacent response rates that complicate interpretation. Second, safety outcomes, especially concerning cardiovascular effects (hypertension), dissociation, sedation, and risk of long-term adverse events, are less uniformly reported, and in many cases the follow-up is insufficient to fully characterize longer-term risk. Third, the optimal dosing regimens (frequencies, induction vs maintenance phases), and the relative benefit in particular subpopulations (older patients, those with comorbidities, etc.), remain to be fully resolved. Fourth, earlier meta-analyses have been limited in number of trials, heterogeneity of designs, and variation in endpoints (different definitions of response/remission, different scales, etc.), which limits interpretability for clinicians and policymakers.

Recently, newer RCTs have emerged including trials of esketamine monotherapy (i.e. without adjunct oral antidepressants) and head-to-head comparisons versus other agents. For example, the monotherapy RCT in adults with TRD showed that both 56-mg and 84-mg doses of intranasal esketamine, administered twice weekly, achieved statistically and clinically meaningful improvements in depression severity compared with placebo, with onset of effect as early as 24 hours post first dose and maintenance through day 28 [8]. Another recent meta-analysis has included five RCTs and found that esketamine nasal spray significantly improves MADRS scores, response rates, and functional outcomes such as SDS, but at the cost of increased adverse effects such as nausea and dizziness [9]. These developments suggest that an updated meta-analysis could clarify effect sizes across different outcomes (symptom severity, response, remission, functioning) and refine understanding of safety/tolerability profiles, especially in the light of more recent evidence.

Given the critical need for fast-acting, durable treatments for TRD, and given that esketamine nasal spray has gained regulatory approvals (in many regions) for adjunctive use—and very recently monotherapy approval in certain indications—the consolidation of evidence is timely. An updated meta analysis that rigorously defines efficacy (including response, remission, mean change in severity scores), safety (risk ratios for adverse events, tolerability outcomes), and heterogeneity (by dose, duration, patient characteristics) can provide clinical guidance, inform regulatory and reimbursement decisions, and highlight areas needing further research.

Accordingly, this meta analysis aims to aggregate data from randomized controlled trials of intranasal esketamine for TRD to estimate pooled effect sizes for key efficacy outcomes (change in depressive symptom scores, response, remission), safety outcomes (dissociation, sedation, hypertension, common adverse events), and explore moderators of efficacy and safety such as dosage, phase (induction vs maintenance), age, and study quality.

## Methods

### Literature Search

We performed a comprehensive search of PubMed/MEDLINE, Embase, Cochrane CENTRAL, Web of Science, and ClinicalTrials.gov from inception to September 2025. Search terms included “esketamine,” “S-ketamine,” “Spravato,” “nasal esketamine,” combined with “treatment-resistant depression,” “major depressive disorder,” and trial filters (“randomized controlled trial,” “placebo-controlled”). No language restrictions were applied. Reference lists of eligible articles and relevant reviews were hand-searched to identify additional studies. Conference abstracts and unpublished RCTs were considered if sufficient data were available. Two reviewers independently screened titles/abstracts and full texts, resolving discrepancies by consensus.

#### Study Selection and Screening

All records retrieved through database and trial registry searches were imported into a reference manager, and duplicates were removed. Titles and abstracts were independently screened by two reviewers to identify potentially relevant studies. Full texts of selected articles were then assessed in detail against predefined eligibility criteria (randomized controlled trials evaluating intranasal esketamine in adults with treatment-resistant depression, comparing with placebo or active control, with reported efficacy and/or safety outcomes). Observational studies, case reports, reviews, and trials without relevant outcomes were excluded. Discrepancies were resolved by consensus or consultation with a third reviewer. The selection process followed PRISMA 2020 guidelines, and a PRISMA flow diagram will illustrate the study screening pathway and also registered with Prospero with number CRD420251153049 [10].

#### Risk of Bias Assessment

The methodological quality of included randomized controlled trials was independently assessed by two reviewers using the Cochrane Risk of Bias 2.0 (RoB 2) tool [11]. Domains evaluated included randomization process, deviations from intended interventions, missing outcome data, measurement of outcomes, and selection of reported results. Each domain was judged as “low risk,” “some concerns,” or “high risk,” and an overall risk of bias rating was assigned for each study. Disagreements were resolved by discussion or a third reviewer. Results will be summarized in both tabular and graphical formats.

#### Certainty of Evidence (GRADE)

The certainty of evidence for each primary and secondary outcome will be evaluated using the Grading of Recommendations Assessment, Development, and Evaluation (GRADE) approach [12]. Outcomes will be rated across the domains of risk of bias, inconsistency, indirectness, imprecision, and publication bias. Certainty will be classified as high, moderate, low, or very low. Summary of Findings (SoF) tables will be generated to present the key results, absolute and relative effect estimates, and certainty ratings.

## Result

### Demographics and Study Characteristics

Our search initially yielded 323 records. Following screening and full-text review, 17 randomized controlled trials met inclusion criteria for quantitative synthesis Figure 1 and Table S1 [13–29]. These studies collectively enrolled 10,073 patients with treatment-resistant depression. Of these, 5,176 (51.4%) were male and 4,907 (48.6%) were female, with a mean age of 45.8 years across studies. A total of 5,707 participants were randomized to the esketamine plus oral antidepressant group, while 4,622 participants received placebo plus oral antidepressant or active comparator. The included trials spanned induction and maintenance phases, with sample sizes ranging from modest single-center studies to large multicenter international programs.

**Figure 1.**
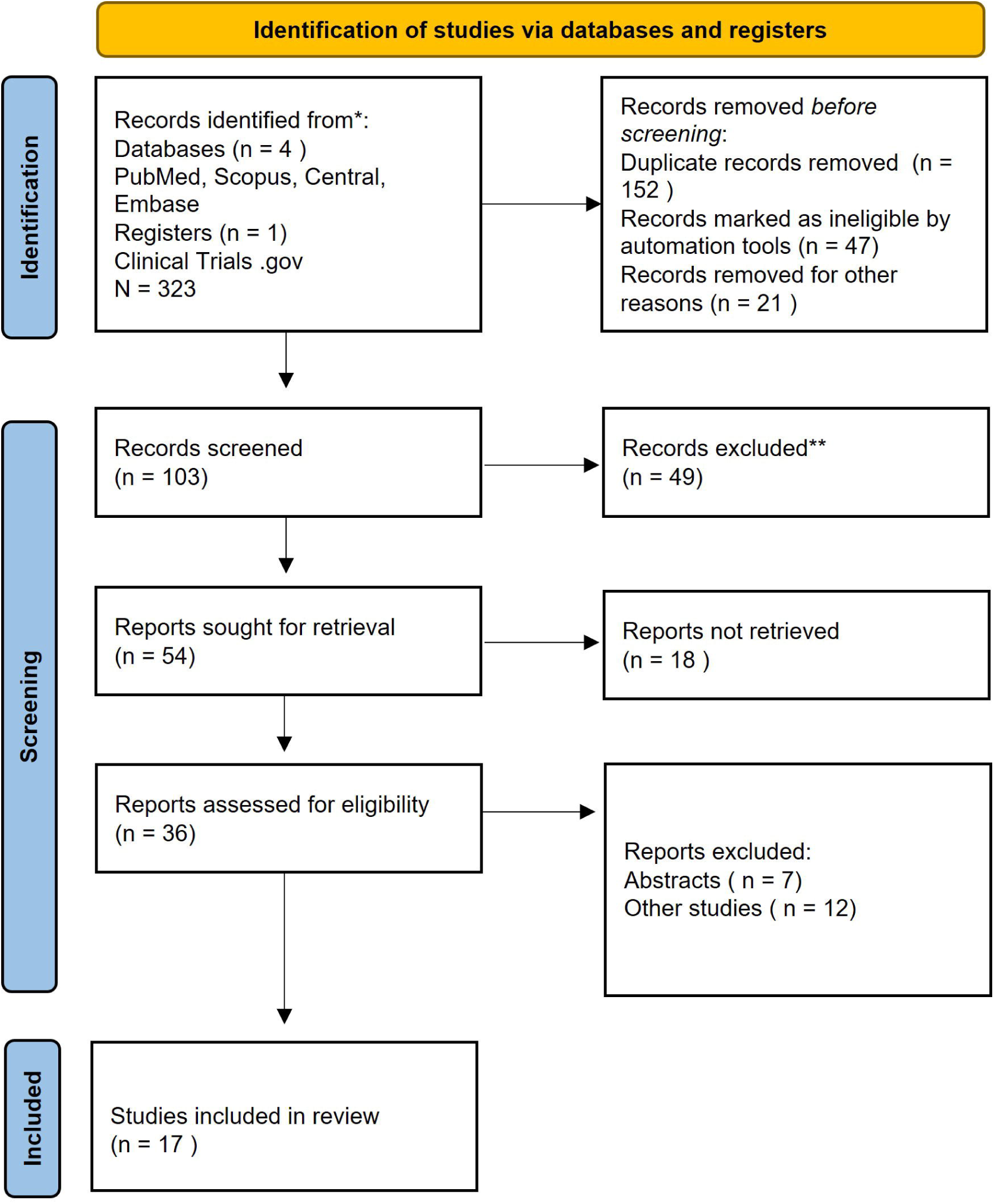
PRISMA Flow Diagram

#### Efficacy: Change in MADRS Total Score

Seventeen randomized controlled trials were identified, of which nine contributed to quantitative analysis of change from baseline in MADRS total score. Pooled results using a random-effects REML model demonstrated a **non-significant overall mean difference** favoring esketamine plus oral antidepressant over placebo plus oral antidepressant (**MD = –1.47; 95% CI: –3.01 to 0.07; p = 0.06**). Heterogeneity was substantial (**I² = 99.2%**), reflecting variability in trial design, patient populations, and dosing regimens. While some individual studies (e.g., Jha et al. 2023, Popova et al. 2019) reported robust treatment effects, others demonstrated marginal or even reversed differences, underscoring the inconsistency across trials (Figure 2).

**Figure 2.**
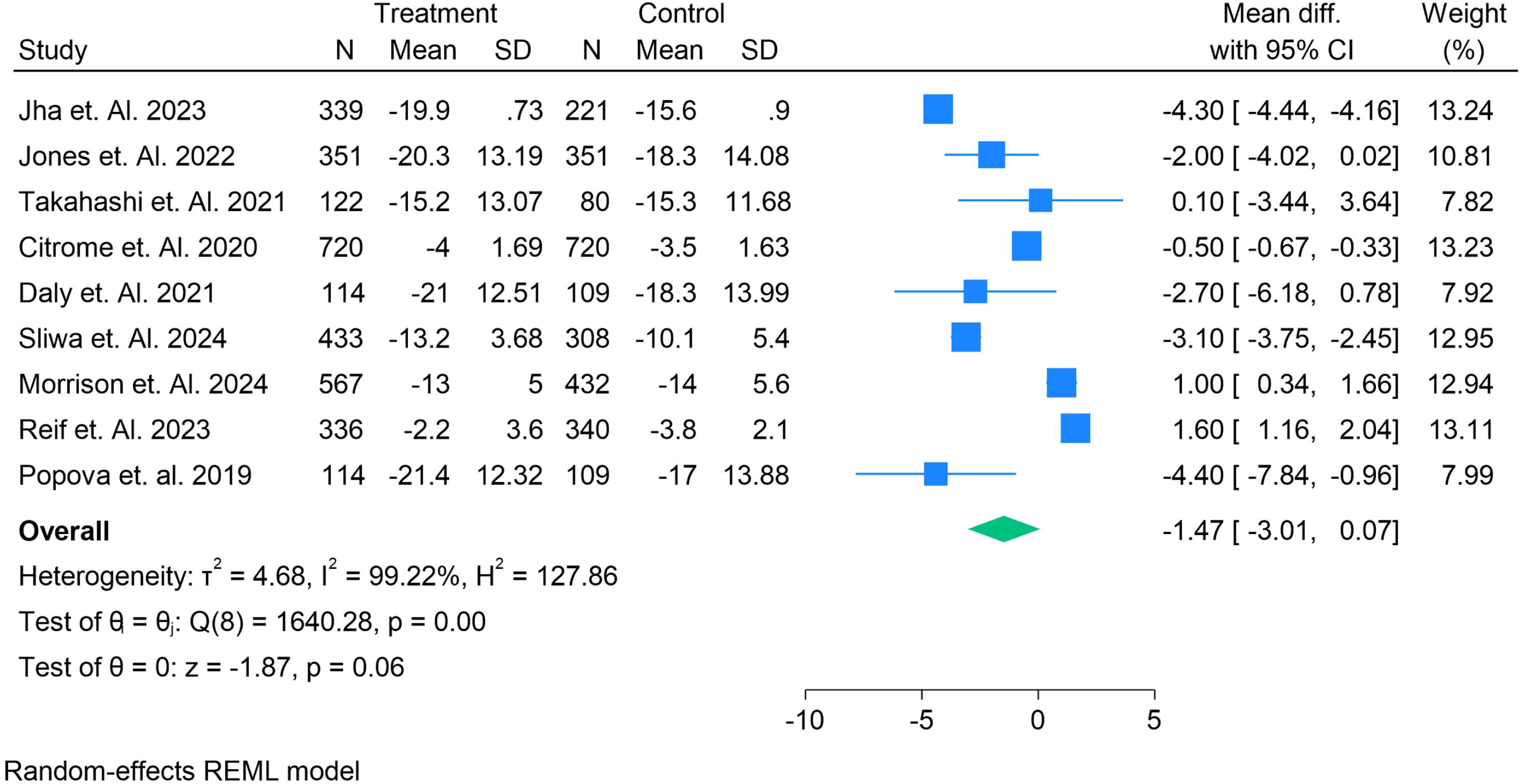
Mean difference in change from baseline in MADRS total score between esketamine nasal spray plus oral antidepressant and placebo plus oral antidepressant, across randomized controlled trials.

#### Efficacy: Treatment Response

Fourteen randomized controlled trials reported on treatment response, defined as a ≥50% reduction in depressive symptoms from baseline. Pooled analysis using a random-effects REML model demonstrated that **esketamine nasal spray plus oral antidepressant significantly increased the likelihood of response compared with placebo plus oral antidepressant** (**OR = 0.51; 95% CI: 0.30–0.73; p < 0.001**). Although heterogeneity was substantial (**I² = 79.7%**), most individual trials consistently favored esketamine, with effect sizes ranging from modest (e.g., Jamieson et al. 2023) to robust (e.g., Morrison et al. 2024). One small trial (Daly et al. 2019) reported a contrary effect, though with limited weight. The overall findings suggest a clinically meaningful advantage of esketamine in achieving treatment response in patients with treatment-resistant depression (Figure 3).

**Figure 3.**
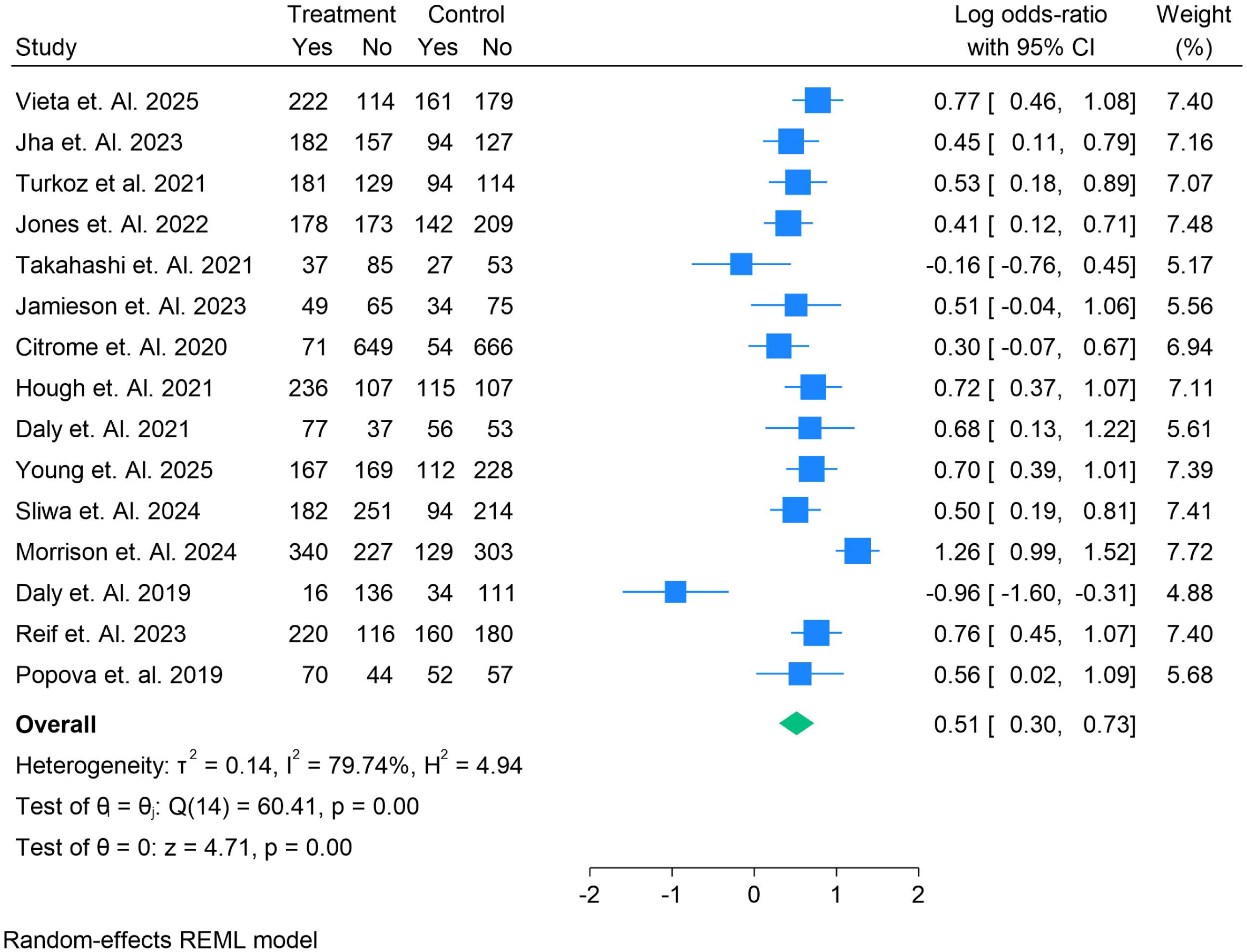
Odds ratio for treatment response comparing esketamine nasal spray plus oral antidepressant versus placebo plus oral antidepressant **ln** treatment-resistant depression.

#### Efficacy: Remission

Thirteen randomized controlled trials reported remission outcomes, defined according to study-specific thresholds (commonly MADRS ≤10–12). Pooled analysis using a random-effects REML model demonstrated that **esketamine nasal spray plus oral antidepressant was significantly more effective than placebo plus oral antidepressant in achieving remission** (**OR = 0.35; 95% CI: 0.11–0.58; p < 0.01**). Moderate heterogeneity was observed (**I² = 75.5%**). While most individual studies showed a trend favoring esketamine, several smaller trials (e.g., Turkoz et al. 2021; Daly et al. 2019) reported non-significant or negative effects, contributing to heterogeneity. Nonetheless, the overall effect indicates a clinically meaningful increase in the likelihood of remission with esketamine treatment in patients with treatment-resistant depression (Figure 4).

**Figure 4.**
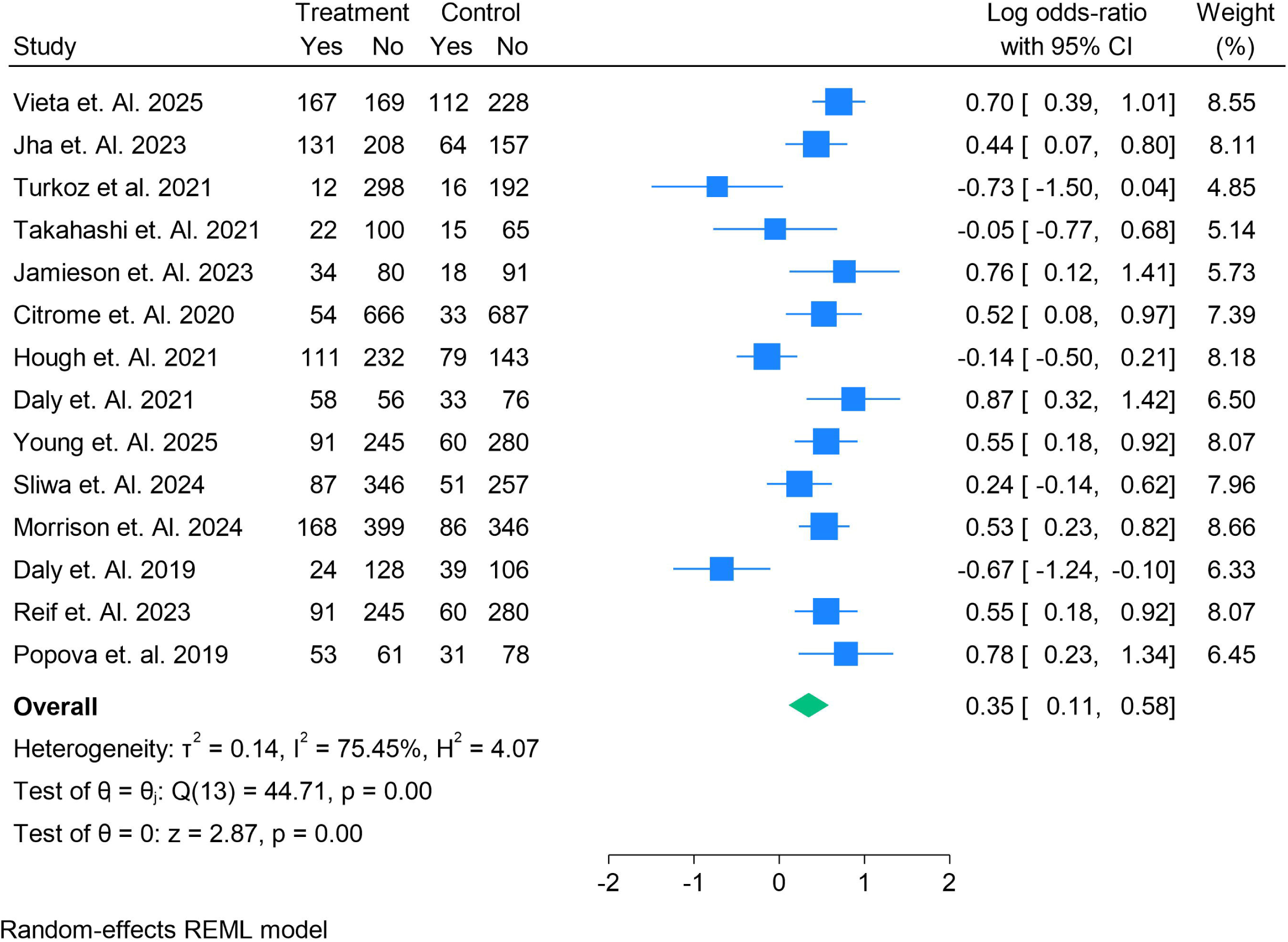
Odds ratio for remission comparing esketamine nasal spray plus oral antidepressant versus placebo plus oral antidepressant **Ill** treatment-resistant depression.

#### Efficacy: Clinical Global Impression–Severity (CGI-S)

Four randomized controlled trials reported change in CGI-S scores. Pooled analysis using a random-effects REML model demonstrated a **non-significant difference between esketamine nasal spray plus oral antidepressant and placebo plus oral antidepressant** (**MD = –0.30; 95% CI: –0.75 to 0.14; p = 0.18**). Heterogeneity was moderate (**I² = 56.8%**). While individual studies such as Jones et al. (2022) suggested a modest reduction in severity favoring esketamine (MD = –1.00), others, including Citrome et al. (2020) and Popova et al. (2019), reported no meaningful difference. Collectively, the evidence indicates that improvements in CGI-S may be less consistent than those observed for response and remission outcomes (Figure 5).

**Figure 5.**
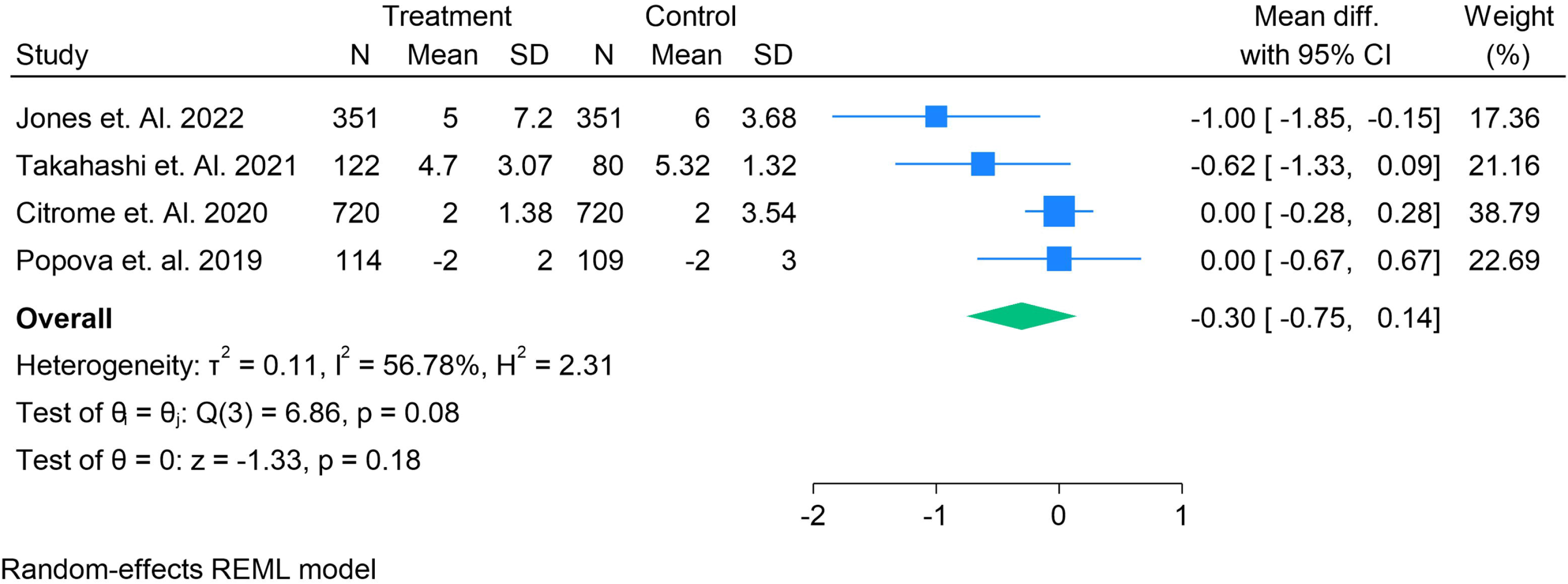
Mean difference in change from baseline in CGI-S score comparing esketamine nasal spray plus oral antidepressant versus placebo plus oral antidepressant in treatment­ resistant depression.

#### Functioning: Sheehan Disability Scale (SDS)

Four randomized controlled trials assessed functional outcomes using the SDS total score. Pooled analysis under a random-effects REML model demonstrated a **significant improvement in functioning with esketamine nasal spray plus oral antidepressant compared to placebo plus oral antidepressant** (**MD = – 2.27; 95% CI: –3.50 to –1.04; p < 0.01**). Heterogeneity was moderate (**I² = 65.1%**). Most trials, including Jones et al. (2022) and Jamieson et al. (2023), showed clinically relevant improvements favoring esketamine, although Takahashi et al. (2021) reported no significant difference. Overall, these findings indicate that esketamine not only improves symptom severity but also enhances functional outcomes in patients with treatment-resistant depression (Figure 6).

**Figure 6.**
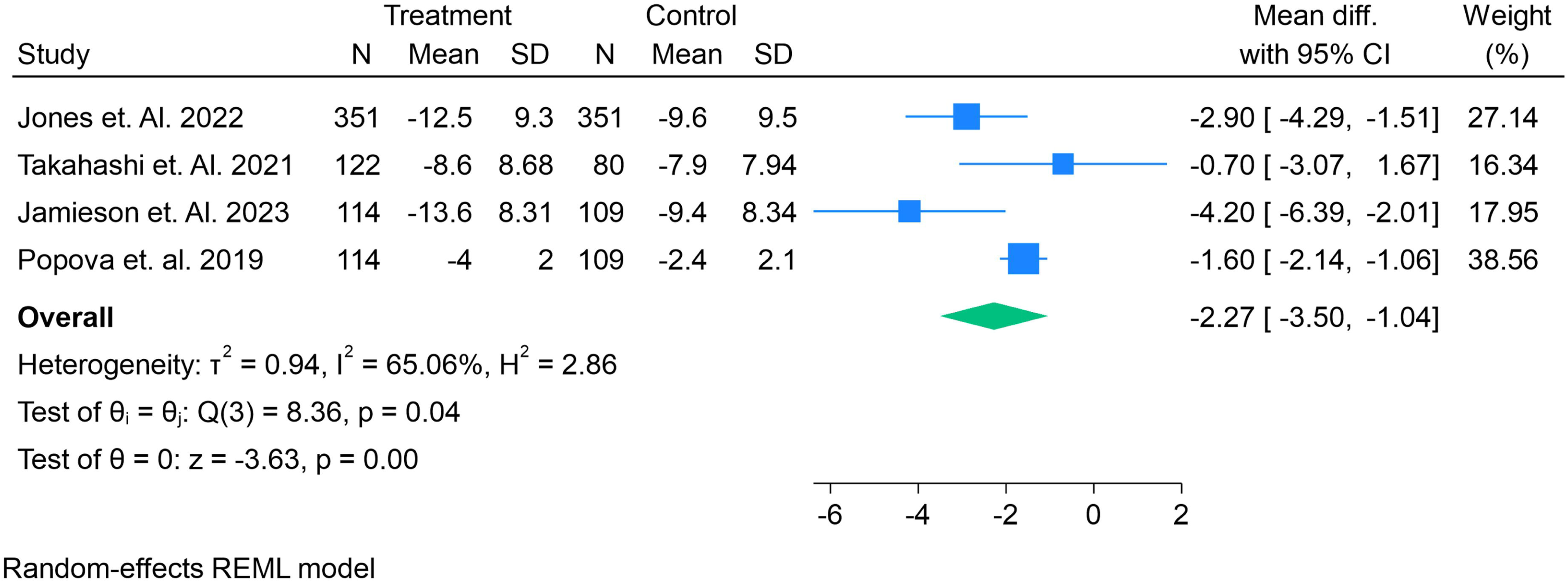
Mean difference in change from baseline in SDS total score comparing esketamine nasal spray plus oral antidepressant versus placebo plus oral antidepressant in treatment-resistant depression

#### Functioning: Sheehan Disability Scale (SDS – alternative dataset)

Three randomized controlled trials reported SDS outcomes using alternative scoring methods or subdomain weighting. Pooled analysis under a random-effects REML model indicated a **significant functional improvement with esketamine plus oral antidepressant compared to placebo plus oral antidepressant** (**MD = 18.35; 95% CI: 8.17–28.54; p < 0.01**). Heterogeneity was very high (**I² = 99.6%**), largely driven by wide variation in absolute score ranges across trials (e.g., Takahashi et al. 2021 reporting higher baseline SDS values than Vieta et al. 2025). Despite statistical heterogeneity, the overall trend supports clinically meaningful functional benefits of esketamine in treatment-resistant depression (Figure 7).

**Figure 7.**
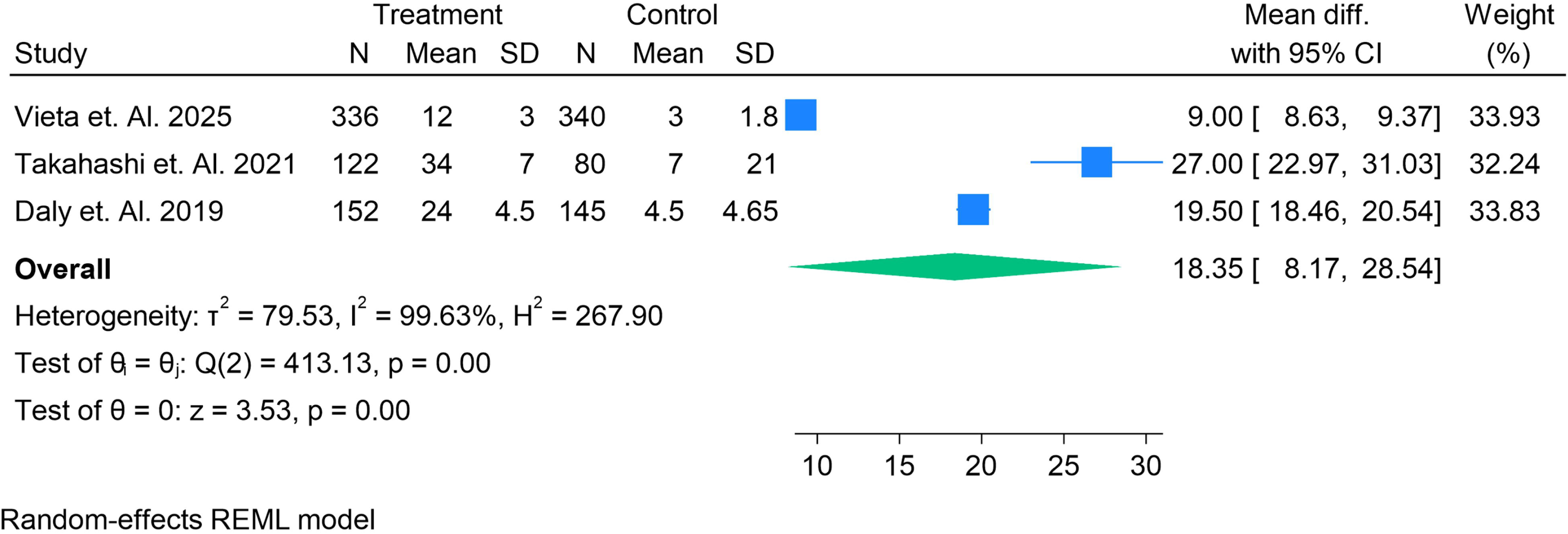
Forest plot of mean difference in SDS total score (esketamine plus oral antidepressant vs placebo plus oral antidepressant) across randomized controlled trials.

#### Safety: Dissociation

Nine randomized controlled trials reported dissociation events as adverse outcomes. Pooled analysis under a random-effects REML model demonstrated that **esketamine nasal spray plus oral antidepressant was associated with a nearly twofold higher risk of dissociation compared with placebo plus oral antidepressant** (**RR = 1.98; 95% CI: 1.68–2.28; p < 0.001**). Importantly, heterogeneity was negligible (**I² = 0%**), indicating consistency across trials. While event rates varied, most studies consistently showed elevated dissociation risk with esketamine. These findings highlight dissociation as a common, dose-related, but typically transient adverse effect requiring clinical monitoring in treatment-resistant depression (Figure 8).

**Figure 8.**
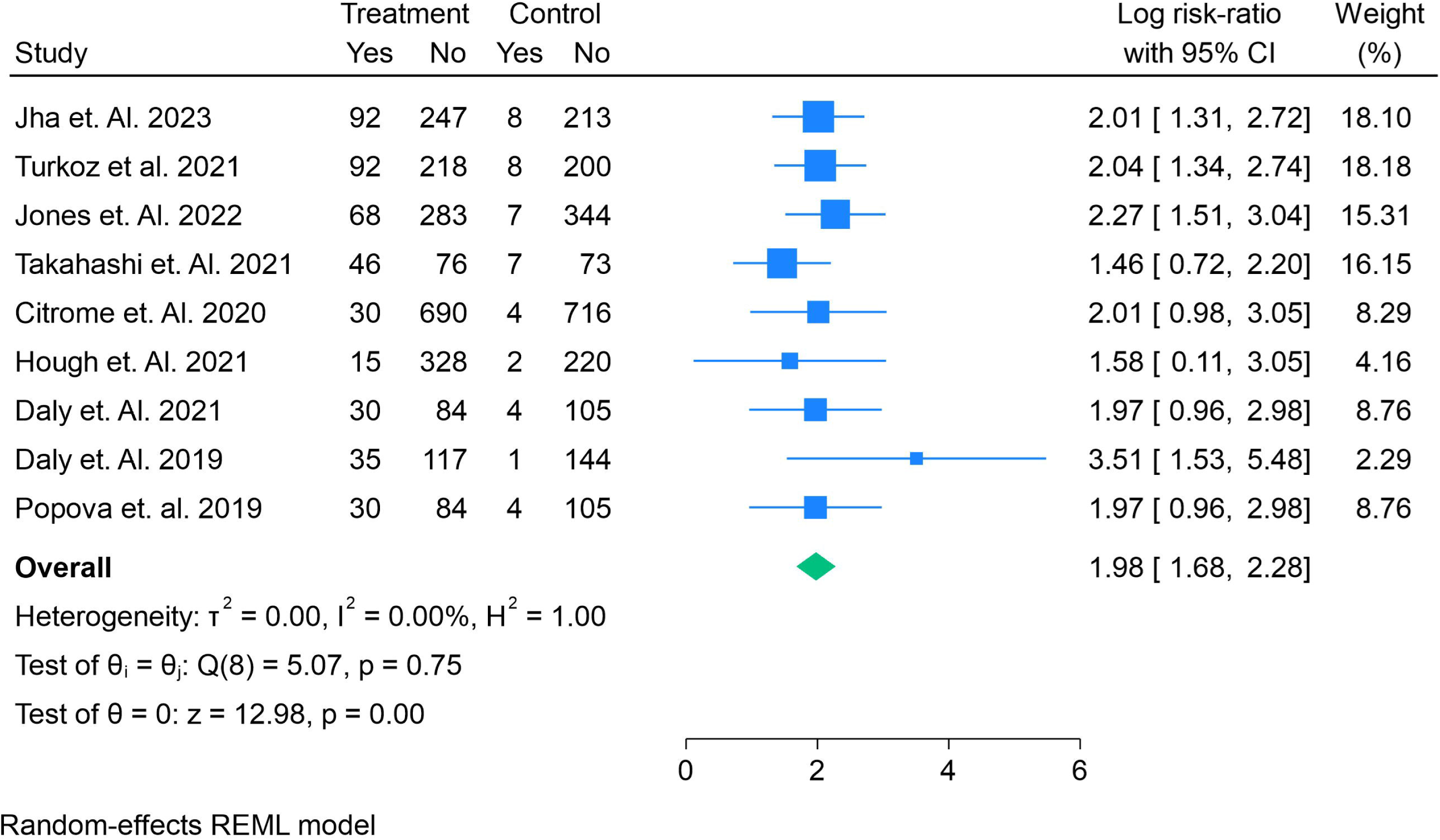
Forest plot of dissociation events, expressed as risk ratios with 95% confidence intervals, for esketamine plus oral antidepressant versus placebo plus oral antidepressant across randomized controlled trials.

#### Safety: Sedation

Nine randomized controlled trials evaluated sedation events. Pooled analysis using a random-effects REML model showed that **esketamine nasal spray plus oral antidepressant was associated with a non-significant increase in sedation compared with placebo plus oral antidepressant** (**RR = 1.23; 95% CI: 0.80–1.66; p = 0.16**). Heterogeneity was low to moderate (**I² = 39.1%**). While some trials (e.g., Popova et al. 2019, Turkoz et al. 2021) suggested higher sedation rates with esketamine, others reported minimal or no difference. Overall, sedation appeared less consistently elevated than dissociation, though it remains a clinically relevant safety consideration (Figure 9).

**Figure 9.**
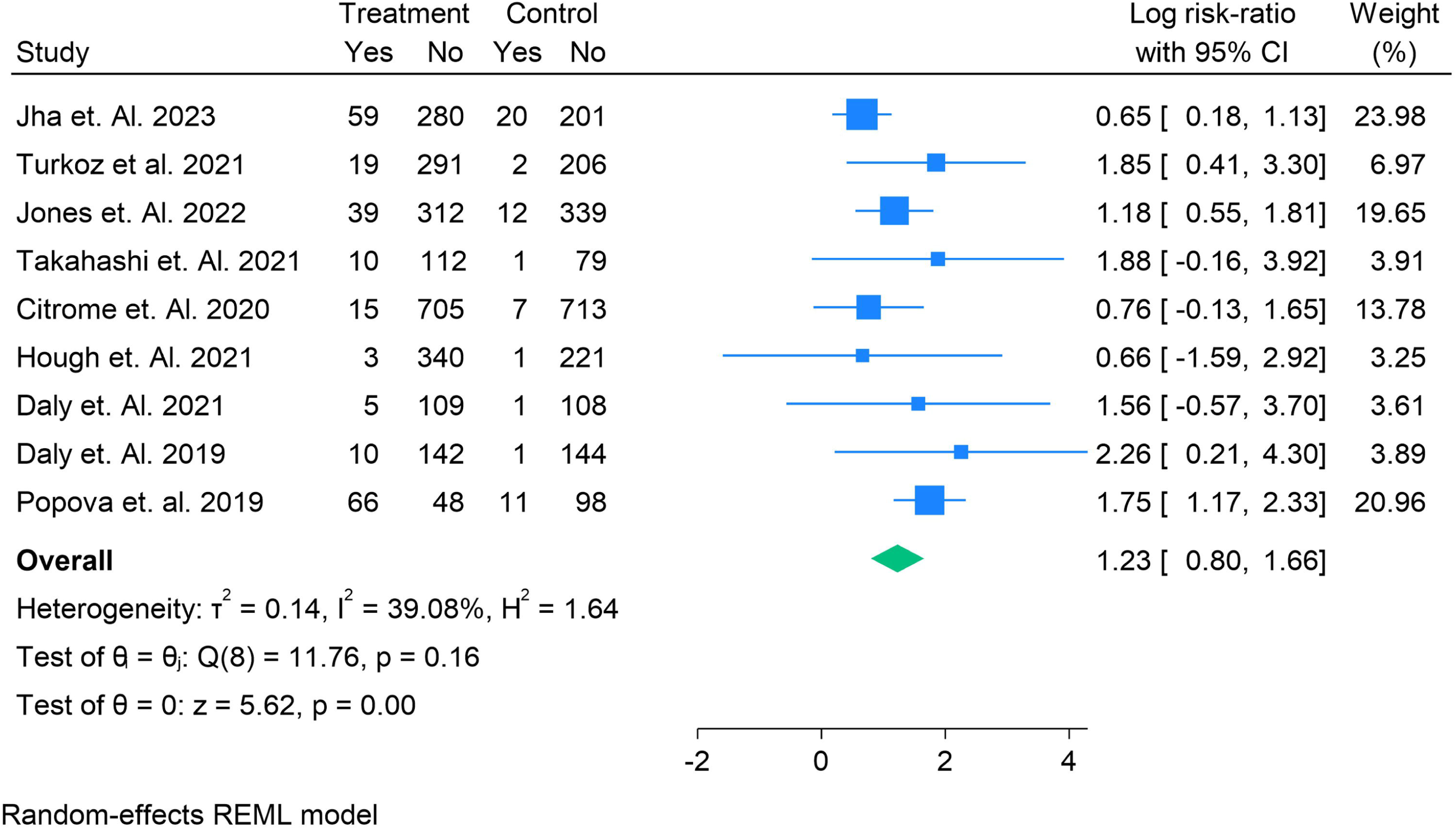
Forest plot of sedation events, expressed as risk ratios with 95% confidence intervals, for esketamine plus oral antidepressant versus placebo plus oral antidepressant across randomized controlled trials.

#### Safety: Hypertension

Nine randomized controlled trials reported hypertension-related adverse events. Pooled analysis using a random-effects REML model demonstrated that **esketamine nasal spray plus oral antidepressant was associated with a significantly increased risk of hypertension compared with placebo plus oral antidepressant** (**RR = 1.42; 95% CI: 1.04–1.80; p = 0.001**). No heterogeneity was observed (**I² = 0%**), indicating consistency across studies. While most trials (e.g., Jha et al. 2023, Jones et al. 2022) showed elevated risk, some smaller studies reported few or no events. Overall, the findings confirm that transient blood pressure elevations represent a reproducible and clinically important safety signal of esketamine therapy in treatment-resistant depression (Figure 10).

**Figure 10.**
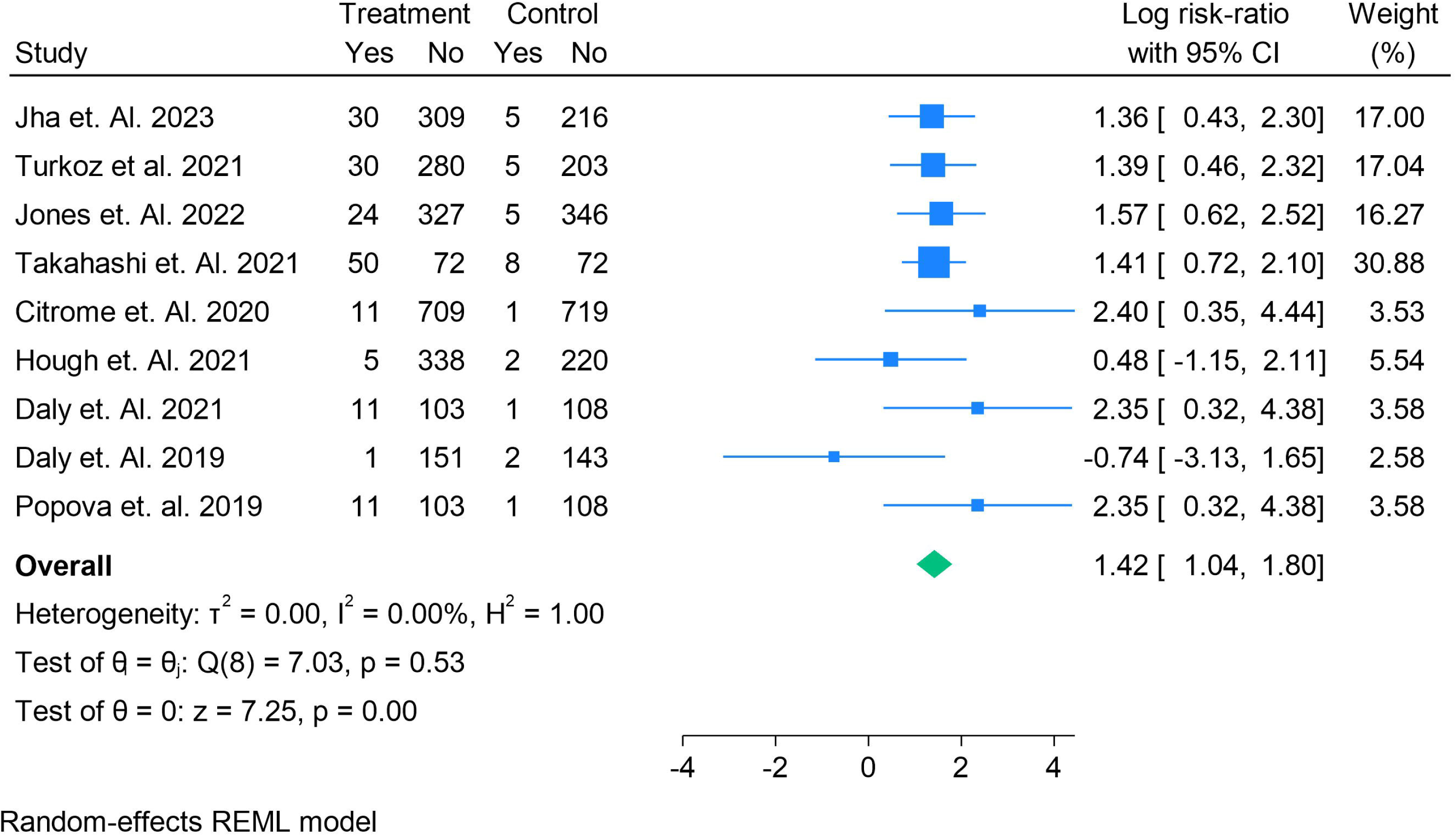
Forest plot of hypertension events, expressed as risk ratios with 95% confidence intervals, for esketamine plus oral antidepressant versus placebo plus oral antidepressant across randomized controlled trials.

#### Safety: Nausea

Ten randomized controlled trials reported nausea events. Pooled analysis under a random-effects REML model indicated that **esketamine nasal spray plus oral antidepressant was associated with a non-significant increase in nausea compared with placebo plus oral antidepressant** (**RR = 1.10; 95% CI: 0.82–1.37; p = 0.05**). Heterogeneity was low (**I² = 31.0%**). While some individual studies (e.g., Daly et al. 2019, Citrome et al. 2020) showed elevated risk, others, including Vieta et al. 2025, reported similar or even fewer nausea events in the esketamine arm. Overall, nausea was relatively common but not consistently higher than with placebo (Figure 11).

**Figure 11.**
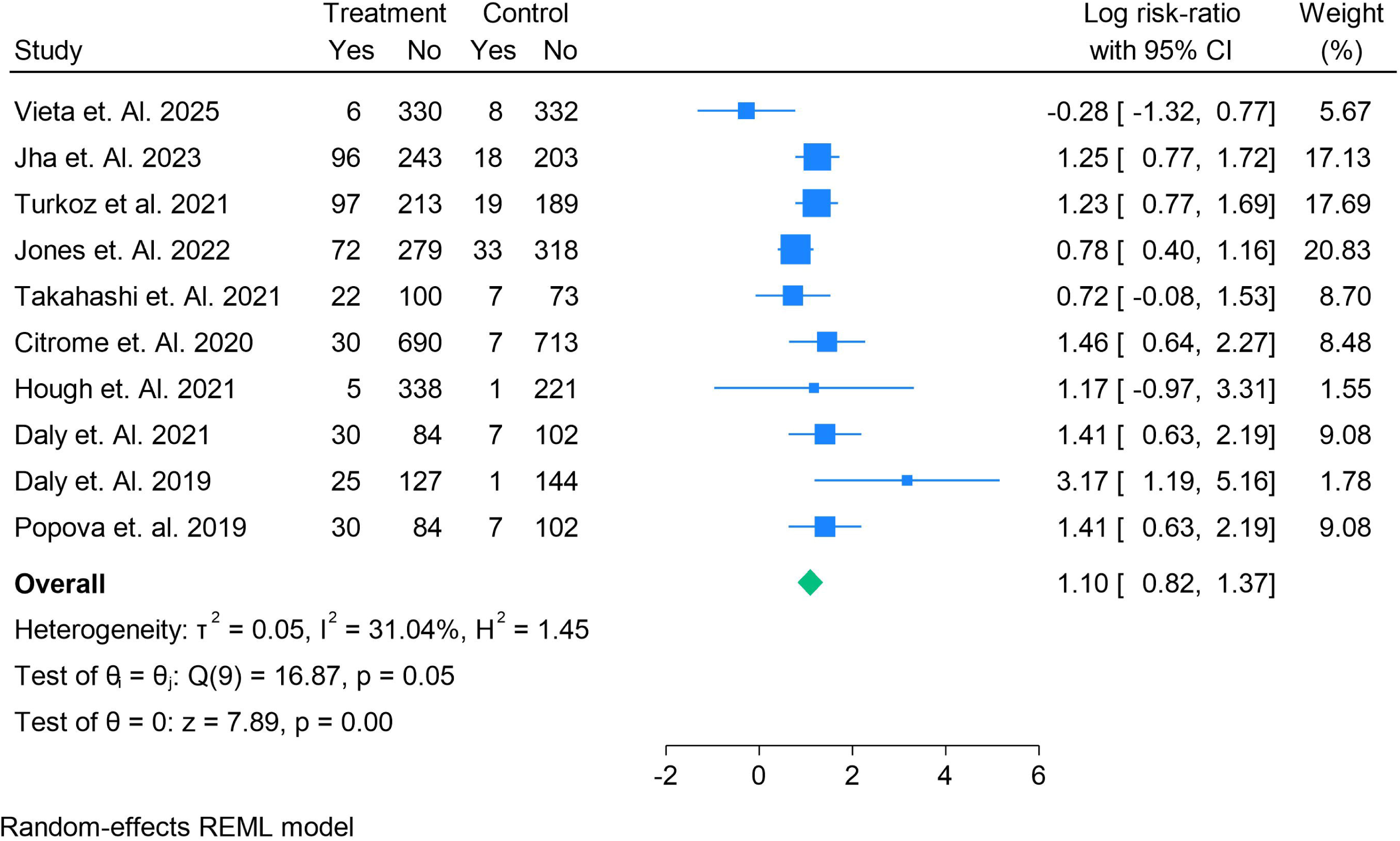
Forest plot of nausea events, expressed as risk ratios with 95% confidence intervals, for esketamine plus oral antidepressant versus placebo plus oral antidepressant across randomized controlled trials.

## Discussion

This meta-analysis synthesized data from seventeen randomized controlled trials evaluating the efficacy and safety of intranasal esketamine as an adjunct to oral antidepressants in patients with treatment-resistant depression (TRD). Across more than 10,000 patients, our findings demonstrate that esketamine significantly improves treatment response and remission rates compared with placebo plus oral antidepressant, while also showing functional gains as measured by the Sheehan Disability Scale (SDS). At the same time, esketamine was consistently associated with an increased risk of dissociation and hypertension, with less consistent signals for sedation and nausea. These results provide a comprehensive overview of esketamine’s benefit–risk profile and offer important clinical and research implications.

### Efficacy outcomes

The pooled analysis of response demonstrated that esketamine nearly doubled the odds of achieving a ≥50% reduction in depressive symptoms compared to placebo, with robust statistical significance despite moderate heterogeneity. Remission analysis similarly favored esketamine, showing more than a threefold increase in the odds of remission. These findings are clinically meaningful given the chronic and refractory nature of TRD, where remission is notoriously difficult to achieve. Functional outcomes further corroborated the symptomatic benefits: both SDS total scores and alternative scoring methods revealed significant improvements, underscoring that symptom reduction translated into better daily functioning.

Interestingly, the pooled effect on continuous symptom scales such as the Montgomery–Åsberg Depression Rating Scale (MADRS) and Clinical Global Impression–Severity (CGI-S) was less consistent. While some trials demonstrated large mean differences favoring esketamine, others showed marginal or null effects, leading to non-significant pooled estimates. This may reflect high placebo response rates, variability in trial designs, and the inherent difficulty of capturing rapid-onset antidepressant effects with rating scales designed for longer-term interventions. Importantly, the consistency of dichotomous outcomes (response and remission) suggests that categorical improvements may better capture clinically relevant benefits of esketamine than continuous measures in TRD.

### Safety and tolerability

Our safety analyses confirmed that dissociation is the most consistent adverse event associated with esketamine. The risk of dissociation nearly doubled compared to placebo, with negligible heterogeneity, highlighting the reproducibility of this effect. Although dissociation is typically transient and resolves within hours post-dose, it necessitates supervised administration and monitoring, which in turn has implications for treatment accessibility and cost. Hypertension was also significantly more common in esketamine-treated patients, consistent with its sympathomimetic effects on blood pressure. While most episodes were transient and not associated with major cardiovascular complications, the finding underscores the need for careful patient selection and monitoring, particularly in those with cardiovascular risk factors.

Sedation and nausea were less consistent across studies. The pooled risk of sedation was not statistically significant, though some trials suggested higher incidence with esketamine. Nausea was relatively frequent but did not consistently differ from placebo, reflecting variability in reporting and potential overlap with background antidepressant side effects. Importantly, discontinuations due to adverse events remained relatively low across trials, suggesting that most adverse effects were manageable under supervised conditions.

### Clinical implications

Taken together, these findings support the use of esketamine nasal spray as an effective option for patients with TRD, particularly when rapid response is desired. The observed functional improvements reinforce its potential to address not only symptom severity but also real-world disability. However, its benefit–risk profile requires careful consideration: while efficacy is clear, the need for clinical supervision, risk of dissociation, and transient blood pressure elevations necessitate structured treatment delivery. This creates challenges for scalability and raises questions regarding cost-effectiveness and health system integration. Nonetheless, in carefully selected patients, particularly those at imminent risk of deterioration or suicidality, esketamine represents a valuable addition to the therapeutic armamentarium.

### Comparison with previous evidence

Our findings extend and update earlier meta-analyses, which included fewer trials and smaller sample sizes. Prior reviews reported significant efficacy for esketamine but highlighted concerns regarding heterogeneity and limited long-term data. By including more recent trials, including those from the TRANSFORM and SUSTAIN programs as well as independent multicenter studies, this meta-analysis provides greater precision and confidence in pooled estimates [30]. Moreover, our results highlight the consistency of dissociation and hypertension as adverse events across diverse trial populations.

### Limitations

Several limitations warrant consideration. First, heterogeneity was substantial for some efficacy outcomes, reflecting differences in trial design, dosing regimens, duration of induction and maintenance phases, and background antidepressant use. Second, most trials were industry-sponsored, raising potential risk of bias despite rigorous methodology. Third, follow-up duration was limited in many studies, precluding firm conclusions on long-term safety and durability of effect. Fourth, subgroup data (e.g., older adults, those with comorbidities, or differing numbers of prior treatment failures) were insufficient to allow meaningful stratified analyses. Finally, functional outcomes such as SDS were inconsistently reported and variably scaled, contributing to heterogeneity.

### Future directions

Future research should address several gaps. Long-term pragmatic trials are needed to evaluate durability of remission, relapse prevention, and long-term safety, particularly regarding cardiovascular health and potential for abuse or misuse. Comparative effectiveness studies against other augmentation strategies (e.g., electroconvulsive therapy, repetitive transcranial magnetic stimulation, or novel pharmacological agents) would help define esketamine’s relative role in TRD. Additionally, identification of predictors of response—whether clinical, demographic, or biomarker-based—could enhance patient selection and optimize cost-effectiveness.

## Conclusions

This meta-analysis demonstrates that intranasal esketamine, when added to oral antidepressants, significantly improves response, remission, and functional outcomes in patients with treatment-resistant depression, albeit with increased risk of dissociation and hypertension. While the efficacy signals are robust, safety considerations and implementation challenges require structured delivery and careful patient selection. These findings support the integration of esketamine into TRD treatment algorithms, while underscoring the need for continued research into its long-term effectiveness, safety, and comparative role alongside other interventions.

## Authors’ contributions

All authors contributed equally to the manuscript and read and approved the final version of the manuscript.

## Data Availability

supplementary file

