## supplementary file for "Efficacy and Safety of Esketamine Nasal Spray in Treatment-Resistant Depression: A Systematic Review and Meta-Analysis of Randomized Controlled Trials"

Supplementary Files

Search String

("esketamine"[Mesh] OR "esketamine"[tiab] OR "S-enantiomer ketamine"[tiab] OR "S-ketamine"[tiab]

OR "JNJ-54135419"[tiab] OR "Spravato"[tiab] OR "intranasal esketamine"[tiab] OR "nasal esketamine"[tiab])

AND

("Depressive Disorder, Treatment-Resistant"[Mesh] OR "treatment-resistant depression"[tiab]

OR "TRD"[tiab] OR "refractory depression"[tiab] OR "major depressive disorder"[tiab]

OR "MDD"[tiab])

AND

("Randomized Controlled Trial"[Publication Type] OR "randomized controlled trial"[tiab]

OR "randomised controlled trial"[tiab] OR "RCT"[tiab] OR "clinical trial"[pt]

OR "double-blind"[tiab] OR "placebo-controlled"[tiab])

Table S1. Findings of all studies

| **Author year** | **Country** | **Total Sample** | **Male** | **Female** | **Age** | **Treatment Name** | **Sample** | **Control Namw** | **Sample** | **Grade** | **Main Finding** |
| --- | --- | --- | --- | --- | --- | --- | --- | --- | --- | --- | --- |
| Vieta et. Al. 2025 | Multicenter | 676 | 229 | 447 | 44.3 | Esketamine nasal spray | 336 | Quetiapine | 340 | High | Esketamine NS plus SSRI/SNRI produced **significantly greater improvements in response, remission, functioning, and workplace productivity** over 32 weeks compared with quetiapine XR plus SSRI/SNRI in patients with treatment-resistant depression |
| Jha et. Al. 2023 | Multicenter | 560 | 392 | 182 | 44.6 | Esketamine nasal spray | 339 | Placebo | 221 | High | Esketamine nasal spray plus oral antidepressant significantly improved depressive symptoms, response, and remission rates compared with antidepressant plus placebo, regardless of baseline irritability, with an acceptable safety profile |
| Turkoz et al. 2021 | Europe | 518 | 404 | 114 | 45.4 | Esketamine nasal spray | 310 | Placebo | 208 | High | Patients with TRD who did not respond within the first week still had a significantly higher likelihood of achieving response at 4 weeks with esketamine nasal spray plus oral antidepressant compared with oral antidepressant plus placebo |
| Jones et. Al. 2022 | USA | 702 | 464 | 238 | 46.6 | Esketamine nasal spray | 351 | Placebo | 351 | High | Esketamine nasal spray plus an oral antidepressant provided **greater improvement in depressive symptoms, response, and remission rates than antidepressant plus placebo**, with a generally similar efficacy and safety profile in both sexes |
| Takahashi et. Al. 2021 | Japan | 202 | 106 | 96 | 45.9 | Esketamine nasal spray | 122 | Placebo | 80 | High | Esketamine nasal spray plus oral antidepressant **was safe and tolerated but did not show statistically significant efficacy over placebo** in Japanese patients with treatment-resistant depression |
| Jamieson et. Al. 2023 | USA | 223 | 112 | 111 | 46.7 | Esketamine nasal spray | 114 | Placebo | 109 | High | *Esketamine nasal spray combined with an oral antidepressant significantly improved health-related quality of life, health status, and functioning in patients with treatment-resistant depression compared with antidepressant plus placebo within 4 weeks* |
| Doty et. Al. 2021 | Multicenter | 1142 | 408 | 734 | 46.3 | Esketamine nasal spray | 855 | Placebo | 432 | High | Esketamine nasal spray, when used with an oral antidepressant, **showed no evidence of adverse effects on olfactory function or nasal health over both short-term and long-term treatment** |
| Citrome et. Al. 2020 | USA | 1440 | 720 | 720 | 48.2 | Esketamine nasal spray | 720 | Placebo | 720 | High | Esketamine nasal spray plus an oral antidepressant improves remission and response rates in treatment-resistant depression, with common but generally manageable adverse events (dissociation, nausea, vertigo) |
| Hough et. Al. 2021 | USA | 565 | 343 | 222 | 54.1 | Esketamine nasal spray | 343 | Placebo | 222 | High | Esketamine nasal spray plus oral antidepressant improves remission and response rates and reduces relapse in treatment-resistant depression, with adverse events that are mostly transient and manageable on the day of dosing. |
| Daly et. Al. 2021 | USA | 223 | 138 | 85 | 44.9 | Esketamine nasal spray | 114 | Placebo | 109 | High | **Esketamine nasal spray plus a new oral antidepressant significantly improved depressive symptoms, response, and remission in treatment-resistant depression regardless of comorbid anxiety, compared with placebo plus antidepressant.** |
| Young et. Al. 2025 | Multicenter | 676 | 338 | 338 | 41.2 | Esketamine nasal spray | 336 | Quetiapine | 340 | High | **Esketamine nasal spray demonstrated consistent and significant superiority over quetiapine XR in achieving remission and preventing relapse in treatment-resistant depression, with robustness across all sensitivity analyses.** |
| Sliwa et. Al. 2024 | USA | 741 | 565 | 176 | 45.7 | Esketamine nasal spray | 433 | Placebo | 308 | High | Esketamine nasal spray + OAD significantly improved depressive symptoms and sustained remission compared to OAD + placebo in treatment-resistant depression |
| Clemens et. Al. 2025 | Bulgaria | 321 | 165 | 156 | 43.2 | Esketamine nasal spray | 165 | Quetiapine | 156 | High | Esketamine nasal spray plus an antidepressant significantly reduced work productivity loss and associated costs compared to quetiapine plus antidepressant in treatment-resistant depression |
| Morrison et. Al. 2024 | USA | 884 | 378 | 506 | 45.2 | Esketamine nasal spray | 567 | Placebo | 432 | High | Esketamine nasal spray plus oral antidepressant did **not negatively impact cognition** in treatment-resistant depression; instead, small improvements in several domains were observed compared with placebo |
| Daly et. Al. 2019 | USA | 297 | 100 | 197 | 45.4 | Esketamine nasal spray | 152 | Placebo | 145 | High | Continuation of esketamine nasal spray plus oral antidepressant significantly delayed relapse in treatment-resistant depression compared with antidepressant plus placebo. |
| Reif et. Al. 2023 | Germany | 676 | 229 | 447 | 47.2 | Esketamine nasal spray | 336 | Placebo | 340 | High | *Esketamine nasal spray plus SSRI/SNRI was superior to quetiapine XR augmentation in treatment-resistant depression, achieving significantly higher remission and response rates with manageable safety profile* |
| Popova et. al. 2019 | Multicenter | 227 | 85 | 138 | 45.2 | Esketamine nasal spray | 114 | Placebo | 109 | High | Esketamine nasal spray plus a newly initiated oral antidepressant produced **significantly greater and faster improvement in depressive symptoms** than oral antidepressant plus placebo in treatment-resistant depression, with manageable transient side effects. |

Figure S1. Risk of Bias

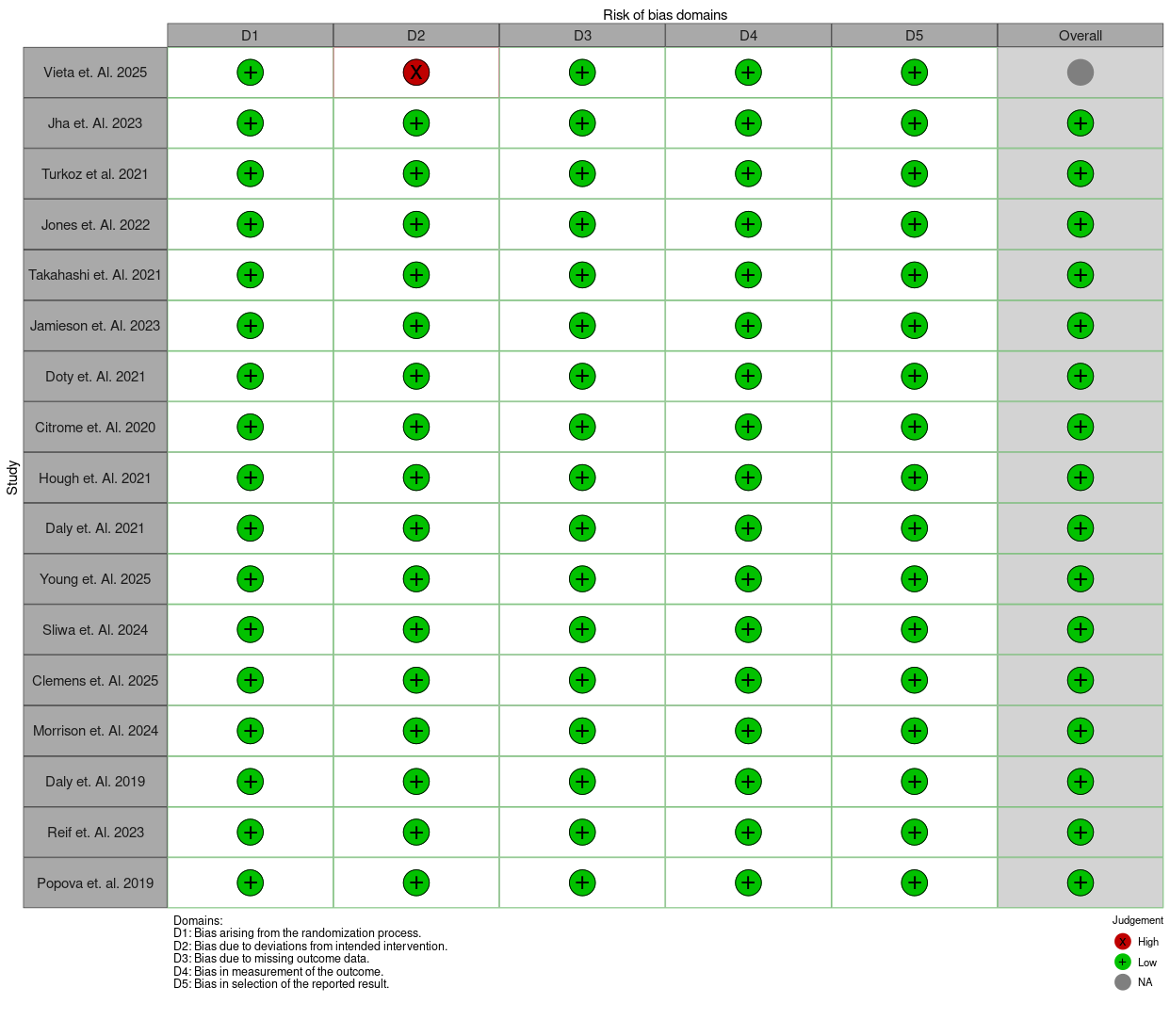
